# The efficacy of vaccines in the context of SARS-CoV-2 and its variants: Role of Spatio-temporal boundary

**DOI:** 10.1101/2021.07.19.21260758

**Authors:** Durbar Roy, Dipshikha Chakravortty, Saptarshi Basu

## Abstract

The preference for a COVID-19 vaccine, among many available, may be difficult for common people and normally relies on efficacy values reported from clinical trials. Vaccine efficacy depends on statistical data from primary and secondary endpoints of a trial. This study provides a time-varying mathematical framework that compares two vaccines of contrasting efficacy (Pfizer-BioNTech and AstraZeneca-Oxford) using hypothetical trials with real-world data. Modeling shows that efficacies can fluctuate depending on the prevailing infection rate and demographics. The efficacy of AstraZeneca-Oxford can become comparable and even better than Pfizer-BioNTech, depending on the region and time of the clinical trials. We also introduce an idea of composite efficacy considering multi-variants and show that the efficacy of various vaccines shows differential sensitivity to the delta variant rampant in India.

## Introduction

COVID-19 (Corona Virus Disease-19) caused by SARS-CoV-2 (Severe acute respiratory syndrome coronavirus 2) virus (*1*) was declared as pandemic on 11th March 2020 by the WHO. The virus caused 188,622,572 positive cases and 4,065,898 deaths worldwide (*2, 3*). The mortality due to COVID-19 occurred mostly in older adults with chronic co-morbid medical conditions. However, a significant share was also present in other age groups. The virus shattered world-wide economies (*4,5*) and health care systems (*6*). Various organizations have undertaken global collaboration and preparedness beyond borders to develop multiple vaccines (*7*) to curtail the disease. These COVID-19 vaccines have undergone various clinical trials to determine their efficacy and safety (*8–10*). Regional and international regulatory authorities decided to approve emergency use of these COVID -19 vaccines for the general public based on the safety and efficacy results. The vaccines (*11*) are primarily characterized and established by determining their safety and efficacy (*12*). The efficacy of a vaccine is defined based on the results obtained from different types of clinical trials (*13–15*). The primary and secondary endpoints (*16*) in the study plan are observed in various kinds of clinical trials. COVID-19 clinical trials involve the inclusion of specific groups of COVID-19 negative human subjects meeting particular inclusion criteria. The subjects have different age ranges, genders, origins, ethnicity with a specific number size K (*17*). The clinical study may have two groups or two arms or more, consisting of human subjects. In a standard clinical trial, the size of each study group is half (K/2) of the total number of subjects recruited. Standard clinical trial ensures test and control groups have at least equal or an adequate number of subjects. The test vaccine is administered in one of the groups. At the same time, the other subgroup is given a placebo (vehicle or formulations without test vaccine). Such studies are generally randomized double blind placebo controlled studies (*18*); i.e., subjects and people who administer vaccines are unaware of the vaccinated and the placebo groups. Then the subjects of both groups are monitored carefully by clinicians for specific clinical study periods ranging from 2-3 months or more. The study period terminates with the estimation of infection rates in the vaccinated (*I*_*v*_) and un-vaccinated groups (*I*_*u*_). The infection rates are calculated from the number of people (percentage) who tested positive for COVID-19 during the study. Further, it is evident from the current COVID-19 pandemic; vaccines are compared based on efficacy data obtained from the various clinical studies of different vaccines. Various government, regulatory authorities, and healthcare organizations have undertaken vaccine-related decisions based on the efficacy values of different vaccines based on the clinical studies (*5*). Robust statistical analysis of clinical trial data would suggest how efficacious the vaccine is for human use. However, there is a critical concern regarding the efficacy of various COVID-19 vaccines reported in the media. The parameters involved in the calculation of efficacy are obtained from clinical trials. A concern exists in the community (*19*) regarding the applicability of clinical trial data for decision-making by ordinary people about the choice of a particular vaccine. Preference for one vaccine over other may also lead to delay in the vaccination of the critical mass if that particular vaccine is not available in sufficient quantity at that time. Emergence of many COVID-19 vaccines in the market implies that the bias against each vaccine needs to be removed so that vaccination by any available vaccine can finally lead to curbing this pandemic promptly. The vaccines, if tested outside standard controlled clinical trials, may lead to variable vaccine efficacy. The efficacy may differ depending on the location, time, prevalent infection rates and the sample size/population (*20*) of the region where the vaccines are administered. Further, the vaccine efficacy may be different against various disease outcomes and also against newly emerging variants of COVID-19 (*21–24*). The vaccine efficacy may also depend on single-dose/complete dose (*25*), double or booster dose, variation on dose intervals, and social distancing measures (*26*). Here, we have explored for the first time the manifestation of the definition of efficacy using a mathematical framework. Literature focusing on the availability of the kind of mathematical analysis presented in this work is relatively sparse (*12*). Hence, this mathematical framework-based knowledge will be essential to elucidate the need for timely vaccination by any available vaccine against COVID-19. This work provides a complete time-varying mathematical framework to understand how vaccine efficacy depends on various statistical quantities obtained from clinical trials. Further, we have provided a framework to calculate the combined vaccine efficacy of COVID-19 vaccines by incorporating the efficacy of the vaccines against the individual variants, especially the delta variant found in India (*27*). Mainly, the study is performed on mRNA BNT162b2 (*19, 28*) and ChAdOx1 nCoV-19 (AZD1222) (*8, 19, 25*) vaccines which are named as Pfizer-BioNTech and AstraZeneca-Oxford respectively. The hypothetical clinical trials for this study are based on real data (*2,3*) from the United States, India, and United Kingdom. Note, however, this analysis is not based on the chemical and biological interactions of the vaccine and virus.

Our study conclusively proves the dependence of vaccine efficacy on geographical location, time of the trial, and the population where the vaccine is being administered. All the vaccines perform equally well in combating viral infection by native or variant strains. Our analysis provides strong evidence for the first time to suggest that no COVID-19 vaccine is superior to others. Every vaccine performs the same based on efficacy values when it comes to protection against infection. Hence, all the vaccines must be given equal preference and timely administration. The emergency use of the vaccine will help the community tackle the disease severity of COVID-19.

## 1 Results (Mathematical framework)

We have carried out the analysis in two parts (1.1 and 1.2). In part 1.1, the distinction between the variants is not present. The distinct variants are lumped into a single variant for which the analysis is presented. Part 2.2 deals with the idea of combined efficacy, where the distinction among different COVID-19 variants is present.

### 1.1 Single variant

The efficacy of any vaccine is defined (*12*) as

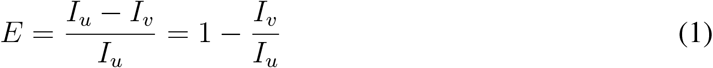

where *I*_*u*_ and *I*_*v*_ are the infection rates of the un-vaccinated and vaccinated people respectively during a clinical trial. The clinical trials are conducted for a specific period at a given region of a population size *K*. From equation (1), it can be observed that the efficacy *E* is a function of two variables *I*_*v*_ and *I*_*u*_. If *I*_*v*_ and *I*_*u*_ are independent quantities then (this assumption is relaxed at a later stage), the changes in efficacy can be calculated using the differential of *E*. Taking the differential of both sides of equation (1) we have

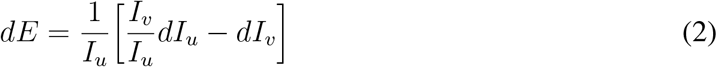

The efficacy sensitivity on *I*_*u*_ and *I*_*v*_ can be calculated using the partial derivatives with respect to the individual variable. The sensitivity of efficacy on *I*_*u*_ is given by

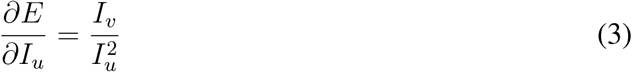

and the sensitivity of efficacy on *I*_*v*_ is given by

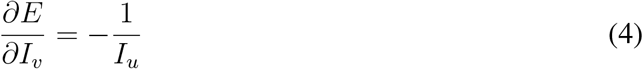

The sensitivity of efficacy *E* on *I*_*u*_ is positive as can be observed from equation (3) and depends on both *I*_*v*_ and *I*_*u*_. However, equation (4) depicts that the sensitivity of *E* on *I*_*v*_ is independent of *I*_*v*_ itself and is a constant negative value (*−*1*/I*_*u*_). For constant *I*_*v*_, the change in efficacy Δ*E* for a given change in Δ*I*_*u*_ can be calculated as

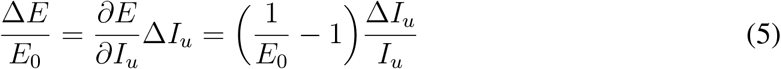

where *E*_0_ is the original efficacy prescribed by the vaccine manufacturers. For constant *I*_*u*_, the change in efficacy Δ*E* for a given change in Δ*I*_*v*_ can be calculated as

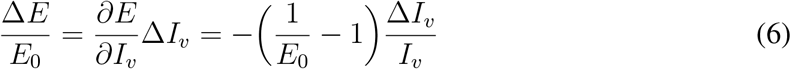

Combining equation (5) and (6), the total change in *E* (Δ*E*) for a given change in *I*_*v*_ (Δ*I*_*v*_) and *I*_*u*_ (Δ*I*_*u*_) is given by

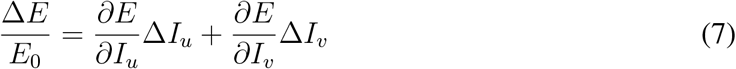

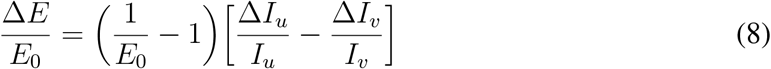

Equation (8) represents the fractional change in efficacy for known fractional changes of *I*_*u*_ and *I*_*v*_, and the original efficacy *E*_0_ provided by the vaccine manufacturers. The analysis described above was based on the assumption that *I*_*v*_ and *I*_*u*_ are independent variables. In the real world, however, a relationship between *I*_*v*_ and *I*_*u*_ exists. *I*_*u*_ is the infection rate in the unvaccinated population. *I*_*u*_ varies similarly to the variation of total active cases in the region where the clinical trials are conducted. A monotonic relationship exists between *I*_*v*_ and *I*_*u*_, i.e., an increase or decrease in *I*_*u*_ causes an increase or decrease in *I*_*v*_ respectively. The positive value of efficacy requires *I*_*v*_ *< I*_*u*_. A wide variation of dependence can be used, however, the most simple dependence is of the form of a power law, given by

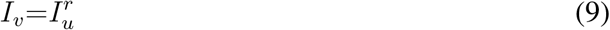

where *r* is a constant evaluated from the initial clinical trial data (conducted by the manufacturers) for individual vaccines. In order to maintain *E >* 0 constraint, *r >* 1 since *I*_*u*_ itself is a proper fraction smaller than unity.

### 1.2 Efficacy in case of multi-variants

The previous section introduced the efficacy of a vaccine in the context of a single virus variant. However, as can be observed during the current COVID-19 pandemic, different variants of SARS-COV-2 have emerged. Clinical trials under such conditions will have an overall efficacy with contributions from individual variants.

Let us develop the combined efficacy during a clinical trial having four variants of COVID-19, say *α, β, γ*, and *δ*. The efficacies with respect to individual variants are given by

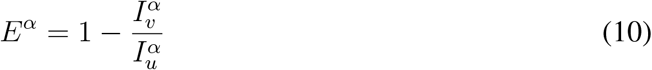

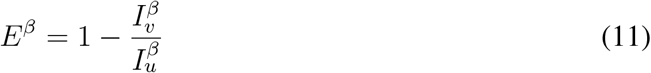

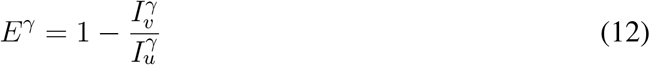

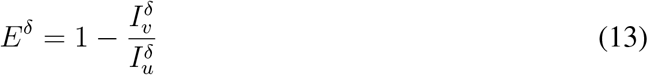

The combined efficacy *E*_*c*_ can be defined using a weighted sum and can be written down in a compact form using the summation notation

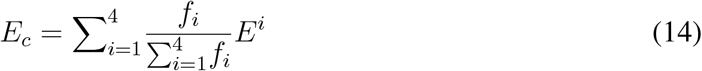

Here *f*_*i*_ represents the efficacy density function for the *i*th variant and acts as a weighting term for *E*^*i*^. For a region having *N* variants, equation (14) can be written as

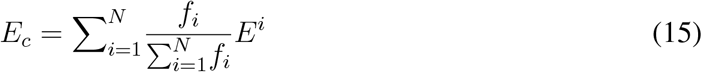

The density function *f*_*i*_ can be thought of as the percentage share of the individual variant. The function *f*_*i*_ depends on time and the region where the clinical trials are performed. Using the percentage share of individual COVID-19 variants as *f*_*i*_ and noting that the sum of percentage shares over all variants will be equal to unity, the denominator of equation (15) will simplify, and the combined efficacy *E*_*c*_ becomes

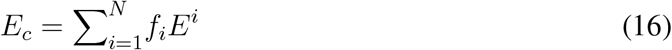

The consequences of the mathematical framework in the context of COVID-19 vaccines is presented in the discussion section.

## 2 Discussion

The efficacy of vaccines is calculated in different clinical trials. It hence depends on the infection rates of the location where the trials are conducted. Further, at a given location, the infection rate itself is evolutionary quantity that change over time. As discussed in the introduction and mathematical framework, more than one variant can affect the efficacy. We will discuss the single variant and multi-variant differently.

### 2.1 Single variant

Fig. 1(A) shows the relative change of efficacy *E* plotted as a function of the relative change of *I*_*u*_ for different initial efficacies *E*_0_ (0.3, 0.5, 0.7, and 0.9) provided by the manufacturers for constant *I*_*v*_. The dependence is linear according to equation (5). The slope of the straight lines is given by the coefficient of the fractional change of *I*_*u*_ in equation (5). An increase in the infection rate *I*_*u*_ increases the efficacy of vaccines provided that *I*_*v*_ remains constant. The invariance of *I*_*v*_ is not apparent in real-world scenarios as an increase of infection rate can increase *I*_*v*_ itself. However, this case can be realized for low infection rates. The assumption of constant *I*_*v*_ is relaxed and discussed at a later stage. The variable nature of infection rates implies that clinical trials performed during different periods of the COVID-19 pandemic will result in different values of efficacies of the same inherent vaccine. For constant *I*_*u*_, the variation of efficacies is shown in Fig. 1(B). An increase in *I*_*v*_ reduces the original efficacy as inferred from the negative values in Fig. 1(B). The trends can be observed from equations (4) and (6). This condition is, however, practically not feasible in the real world. Fig. 1(C) shows the general trend of *I*_*v*_ with respect to *I*_*u*_ for a vaccine to maintain a constant efficacy *E*_0_ (*E*_0_ is a parameter). Fig. 1(D) shows the total number of active COVID-19 cases (in millions) in India at five discrete points in June 2020, September 2020, December 2020, March 2021, and May 2021. If hypothetical clinical trials were performed during periods A, B, C, and D (labeled in Fig. 1(D)) for different vaccines, the efficacy would be different as the infection rate during the four phases is different. The most commonly advocated vaccines of COVID-19 and their corresponding efficacies as prescribed by the manufacturers are shown in Fig. 1(E). Pfizer-BioNTech, Moderna, Sputnik all have efficacies above ninety percent *E*_0_ *>* 0.9, whereas the vaccine provided by AstraZeneca-Oxford has an efficacy of *E*_0_ = 0.67. Fig. 1(F) shows the changes in *I*_*v*_ for a change of *I*_*u*_ during the pandemic for maintaining a constant efficacy. It can be observed that the corresponding swings in *I*_*v*_ are more significant for vaccines having lower efficacies (like AstraZeneca-Oxford) when compared to vaccines having higher efficacies (like Pfizer-BioNTech, Moderna, and Sputnik) for a given change in *I*_*u*_. Table 1 shows the exact values of Δ*I*_*v*_ for different values of Δ*I*_*u*_ for four different vaccines in order to maintain a constant efficacy as prescribed by the manufacturers. In contrast to constant efficacy, constant *I*_*v*_ can model many actual clinical trials and is essential in real-world scenarios. In general, the efficacy of a vaccine will change as the pandemic progresses in time. Table 2 tabulates the relative change of efficacies of four different vaccines as a function of the relative change in *I*_*u*_. In general, *I*_*u*_ cannot be controlled and is a function of the local infection rate where the clinical trials are being conducted (i.e., *I*_*u*_ is dependent on the different phases of the pandemic), whereas *I*_*v*_ is a quantity that measures how good a vaccine is in a given region and time. The two different extreme vaccines Pfizer-BioNTech and AstraZeneca-Oxford, have been chosen based on their *E*_0_ values for the remaining analysis and the data (*2, 3*) for hypothetical clinical trials in the United States, India, and United Kingdom. Fig. 2(A) shows the number of active COVID-19 cases as a function of time for different geographical locations (the United States, India, and the United Kingdom). The curves have many distinct peaks showing different waves that different regions experienced during the progress of the pandemic. The blue, red, and green curves represent the United States of America, India, and the United Kingdom. The maximum active cases for these regions are different, and the peaks occur at different time instants. The active cases are approximately a good measure for calculating *I*_*u*_ for the population of a given region. *I*_*v*_ and *I*_*u*_ were initially considered as independent variables; however, in real-world conditions, a dependence exists between *I*_*v*_ and *I*_*u*_. Fig. 2(B) shows the relationship between *I*_*v*_ and *I*_*u*_ in various clinical trials for Pfizer-BioNTech. A similar relationship for AstraZeneca-Oxford is shown in Fig. 2(C). Fig. 2(D) represents *I*_*u*_ for United States of America, India, and United Kingdom. *I*_*u*_ is calculated from a hypothetical clinical trial on the total population size of the regions. The existence of a monotonic relationship between *I*_*v*_ and *I*_*u*_ can be inferred from the clinical data in Fig. 2(B) and 2(C). The clinical trial data is represented according to equation (9) using a least-square analysis (*29*). The exponent of *I*_*u*_ in equation (9) has been extracted for Pfizer-BioNTech and AstraZeneca-Oxford from the least-square fit. The exponent *r* for Pfizer-BioNTech and AstraZeneca-Oxford are 1.6 and 1.25, respectively. Fig. 2(E) shows the temporal variation of *I*_*v*_ for all the countries and the vaccines, calculated using equation (9). Coupling equation (1) and equation (9), Fig. 2(F) shows the variation of the efficacy as a function of time for two different vaccines (Pfizer-BioNTech and AstraZeneca-Oxford) for hypothetical trials conducted in all the countries. As it can be inferred from the plots, the efficacy values, in general, oscillates with time. However, as it can be observed, the curves of Pfizer-BioNTech and AstraZeneca-Oxford intersect each other, indicating similar efficacies at the point of intersection. A similar analysis is carried out on a much more local scale where the sample size (population) is much smaller. Fig. 3(A) shows the active cases in New York (US) and Maharashtra (India) as a function of time. Fig. 3(B) and 3(C) show the temporal variation of *I*_*u*_ and *I*_*v*_, respectively. From Fig. 3(B), it can be inferred that adequate sample size is essential; the maximum local infection rate *I*_*u*_ for New York reaches approximately nine percent. However, on a much larger scale of the entire country of the United States, the maximum value of *I*_*u*_ was about six percent (refer Fig. 2(D)). Hence the vaccine efficacy values of Pfizer-BioNTech and AstraZeneca-Oxford can be similar depending on the region, time, and sample size of the clinical trials.

**Figure 1:**
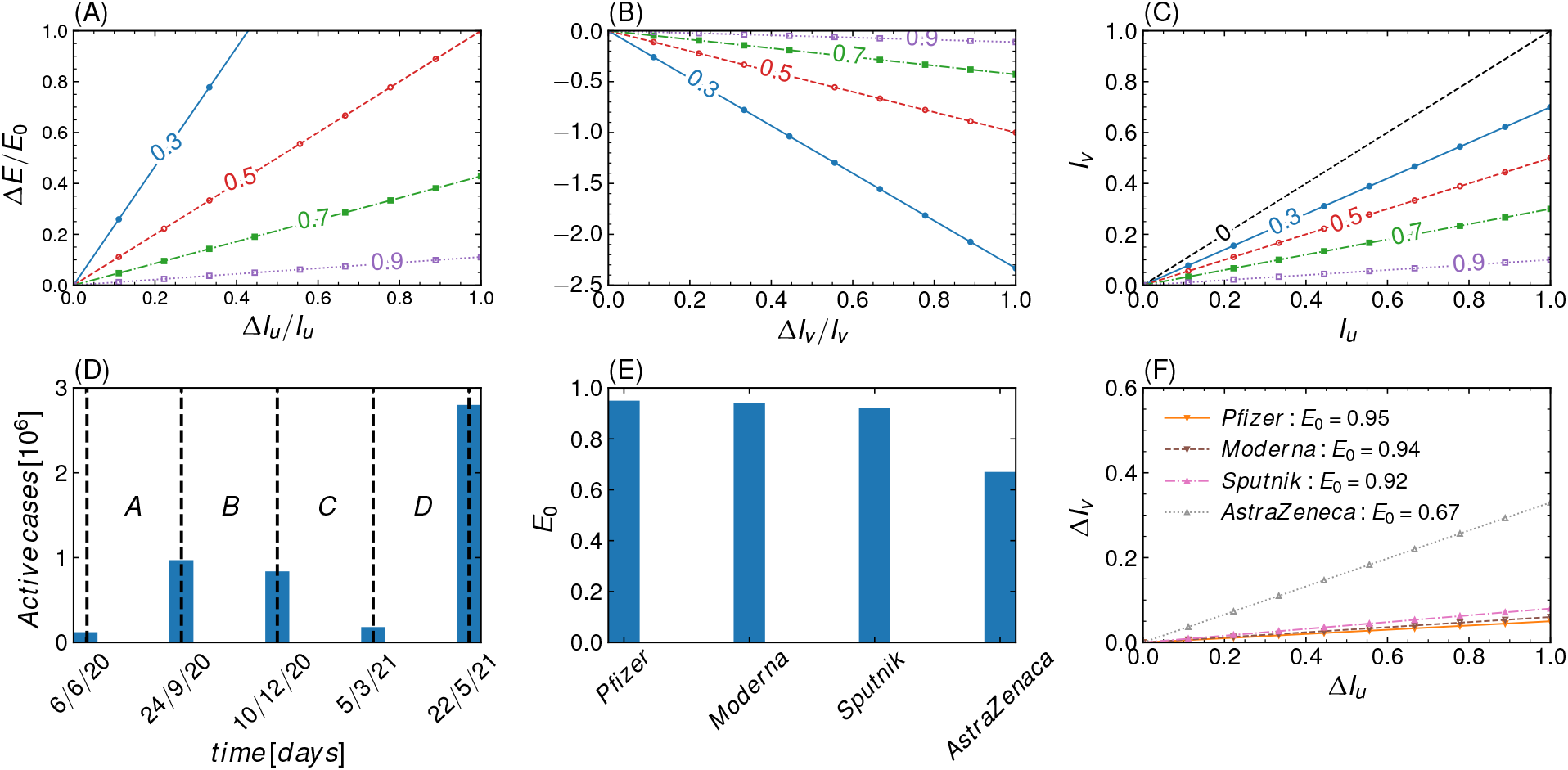
Mathematical implication of equation (1-8). Sensitivity dependence of *E* on *I*_*u*_ and *I*_*v*_. **(A)** The fractional change in *E* plotted as a fractional change of *I*_*u*_ with original efficacy *E*_0_ as a parameter and constant *I*_*v*_. **(B)** The fractional change in *E* plotted as a fractional change of *I*_*v*_ with original efficacy *E*_0_ as a parameter and constant *I*_*u*_. **(C)** *I*_*v*_ plotted as a function of *I*_*u*_ with (constant efficacy) *E*_0_ as a parameter. **(D)** Confirmed COVID-19 cases in millions plotted as some discrete points in time (dates) for India. A, B, C, D represents the period of 4 hypothetical clinical trials. **(E)** Comparison of different COVID-19 vaccines based on their efficacies. **(F)** Δ*I*_*v*_ plotted as a function of Δ*I*_*u*_ if *E* is restricted to change. The curves are plotted for four different COVID-19 vaccines.

**Table 1:** Table of Δ*I*_*v*_ for different vaccines and different Δ*I*_*u*_ values for hypothetical clinical trials based on Fig. 1(D). A, B, C, D represents the period of the clinical trials.

|  | Pfizer-BioNTech | Moderna | Sputnik | AstraZeneca-Oxford |
| --- | --- | --- | --- | --- |
| $E_0$ | 0.95 | 0.94 | 0.92 | 0.67 |
| $\Delta I_u$ | | | | |
| 0.10 | 0.005 | 0.006 | 0.008 | 0.033 |
| 0.30 | 0.015 | 0.018 | 0.024 | 0.099 |
| 0.40 | 0.02 | 0.024 | 0.032 | 0.132 |
| 0.50 | 0.025 | 0.03 | 0.04 | 0.165 |

**Table 2:** Table of Δ*E/E*_0_ for different vaccines and different values of Δ*I*_*u*_*/I*_*u*_ for hypothetical clinical trials based on Fig. 1(D).

|  | Pfizer-BioNTech | Moderna | Sputnik | AstraZeneca-Oxford |
| --- | --- | --- | --- | --- |
| $E_0$ | 0.95 | 0.94 | 0.92 | 0.67 |
| $\Delta I_u/I_u$ | | | | |
| 0.10 | 0.0053 | 0.0064 | 0.0087 | 0.0492 |
| 0.30 | 0.0158 | 0.0191 | 0.0261 | 0.1478 |
| 0.40 | 0.0210 | 0.0255 | 0.0348 | 0.1970 |
| 0.50 | 0.0263 | 0.0319 | 0.0435 | 0.2463 |

**Figure 2:**
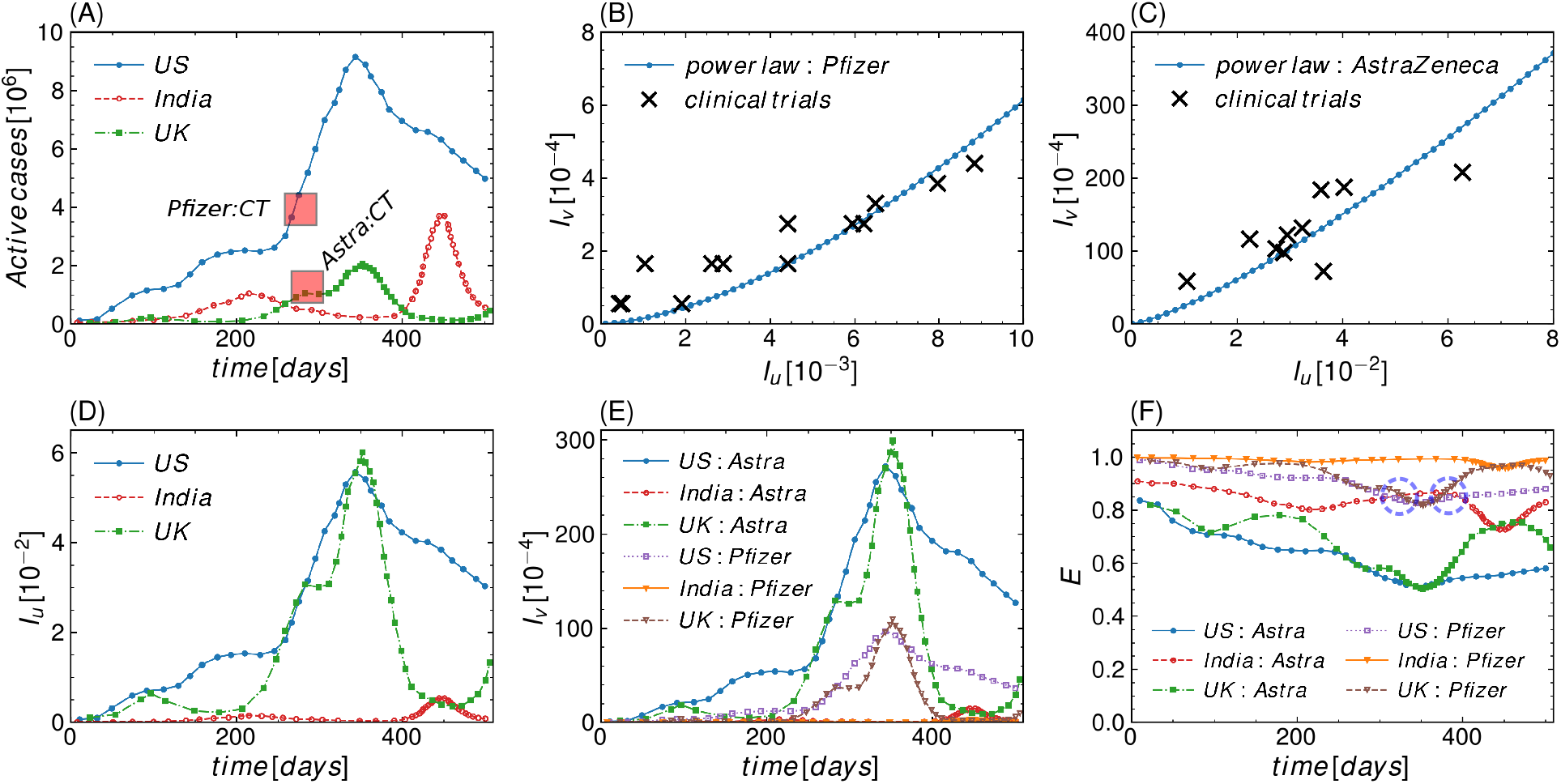
Vaccine efficacy and its parameters for hypothetical clinical trials based on different countries (large sample size). In the figures Pfizer-BioNTech, AstraZeneca-Oxford, United States of America, United Kingdom, Clinical Trials are reffered as Pfizer, Astra, US, UK and CT respectively. **(A)** Active COVID-19 cases in India, United States and United Kingdom plotted as a function of time. The red boxes denotes the time at which the efficacy of Pfizer-BioNTech and AstraZeneca-Oxford was declared (*2, 3*). **(B)** Dependence of *I*_*v*_ on *I*_*u*_ for Pfizer-BioNTech in different clinical trials (*19*) (black cross) and the power law fit (blue curve). **(C)** Dependence of *I*_*v*_ on *I*_*u*_ for AstraZeneca-Oxford in different clinical trials (*8*) (black cross) and the power law fit (blue curve). **(D)** *I*_*u*_ as a function of time for United States, India and United Kingdom. **(E)** *I*_*v*_ (calculated from the power law dependence) as a function of time for United States, India and United Kingdom for both AstraZeneca-Oxford and Pfizer-BioNTech. **(F)** *E* as a function of time for United States, India and United Kingdom for both AstraZeneca-Oxford and Pfizer-BioNTech. The violet dotted circles denotes crossover points during which the efficacy of Pfizer-BioNTech and AstraZeneca-Oxford are the same.

**Figure 3:**
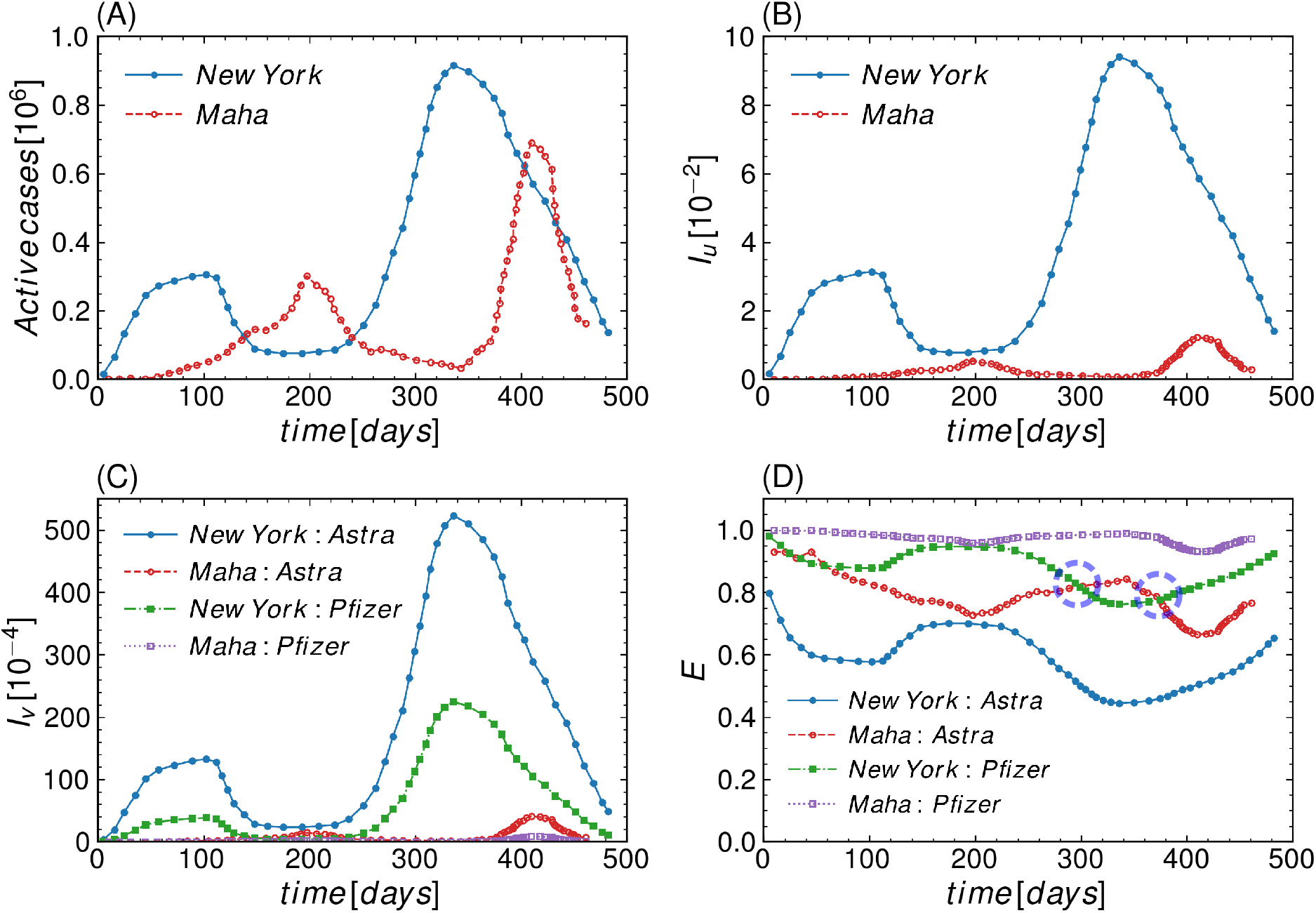
Vaccine efficacy and its parameters for hypothetical clinical trials based on different states of countries (relatively small sample size). In the figures Pfizer-BioNTech, AstraZeneca-Oxford and Maharashtra, are reffered as Pfizer, Astra, Maha respectively. **(A)** Active COVID-19 cases in New York and Maharashtra plotted as a function of time (*2, 3*). **(B)** *I*_*u*_ as a function of time for New York and Maharashtra. **(C)** *I*_*v*_ (calculated from the power law dependence) as a function of time for New York and Maharashtra for both AstraZeneca-Oxford and Pfizer-BioNTech. **(D)** *E* as a function of time for New York and Maharashtra for both AstraZeneca-Oxford and Pfizer-BioNTech. The violet dotted circles denotes crossover points during which the efficacy of Pfizer-BioNTech and AstraZeneca-Oxford are the same.

### 2.2 Multi-variants

As deduced in section 1.2, the distinction between different variants can change the combined efficacy *E*_*c*_. In the presence of multiple variants, the combined efficacy of a vaccine can be evaluated using equation (16) in a given clinical trial. As the COVID-19 pandemic was evolving with time, several variants of COVID-19 were discovered at different places globally like B1.1.306, B1.617.1, and B1.617.2 (*22, 23*) to name a few. The B1.617.2, also called the delta variant, was found in India (*27*) and is thought to be responsible for the second wave in India. In real-world scenarios, several variants can coexist and evolve simultaneously, with time changing the course of the pandemic. Fig. 4(A) shows the percentage share of all the variants during 300 days in India (*24*). It can be observed that each of the individual variants has its own evolutionary transient dynamics. It can be observed how a particular variant becomes dominant (delta variant here) over a certain period (last 60 days in Fig. 4(A)). Pfizer-BioNTech and AstraZeneca-Oxford have reported (*30, 31*) different efficacies for some variants of COVID-19 shown in Fig. 4(B) and 4(C). The individual efficacies (*E*^*i*^) concerning a given variant, along with the percentage share of the variants (*f*_*i*_), are used to calculate the combined efficacy using equation (16). The *f*_*i*_ in equation (16) is the percentage share of the individual variants that evolve with time, resulting in transient variation of the combined efficacy. Suppose the efficacy of a clinical trial is evaluated daily. In that case, the combined efficacy of the vaccine will have an evolutionary nature governed by the dynamics of the individual variants. Fig. 4(D) shows the combined efficacy of Pfizer-BioNTech and AstraZeneca-Oxford over 60 days, where the delta variant rises steeply in India (refer Fig. 4(A)). The combined efficacy of Pfizer-BioNTech is more sensitive to the delta variant (decreases quite steeply) variant than that of AstraZeneca-Oxford. The steepness of the Pfizer-BioNTech curve is due to the combined effect of the sharp increase of the delta variant, decrease of the other variants, and the low efficacy of Pfizer-BioNTech for the delta variant compared to AstraZeneca-Oxford.

**Figure 4:**
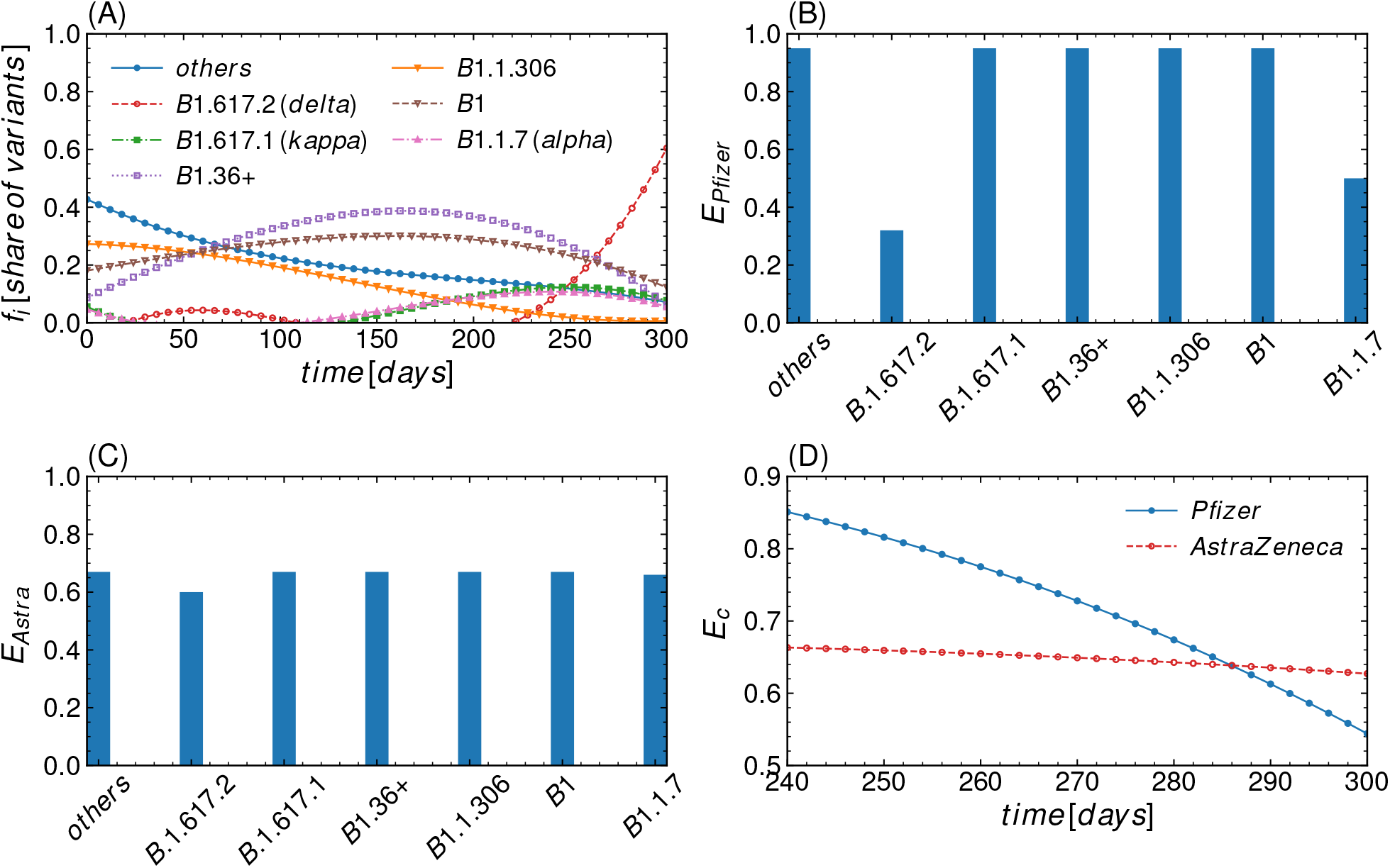
Role of multi-variants on the efficacy of vaccines. In the figures Pfizer-BioNTech, AstraZeneca-Oxford are reffered as Pfizer, Astra respectively. **(A)** The percentage share of different COVID-19 variants in India represented as a function of time in days (*24*). **(B)** Pfizer-BioNTech vaccine efficacies for different individual variants obtained from clinical trials (*32*). **(C)** AstraZeneca-Oxford vaccine efficacies for different individual variants obtained from clinical trials (*31*). **(D)** Comparing combined vaccine efficacies of Pfizer-BioNTech and AstraZeneca-Oxford for different variants.

## 3 Conclusion

In conclusion, we have studied the mathematical definition of efficacy and its implications on statistical quantities like *I*_*v*_ and *I*_*i*_. The efficacy used to characterize a COVID-19 vaccine depends on the infection rate of the region, sample size, and time when the clinical trials are performed. Further, we have also shown that the definition of vaccine efficacy can be generalized by incorporating multi-variants efficacy into consideration. Therefore, the definition of efficacy used by vaccine manufacturers globally is a spatio-temporal quantity that can vary from region to region and evolve with time. Hence the vaccine efficacy defined may not be solely the property of the vaccine itself, i.e., not an inherent property of the vaccine alone. However, it may also depend on the parameter characterizing the pandemic. Hence, efficacy data should be used with utmost care and understanding. The bias towards a better choice of vaccines based on efficacy values is often misleading and requires a thorough reconsideration.

## Data Availability

Data available within the manuscript

https://www.overleaf.com/project/60f3f4972d7eee548b39b610

## Acknowledgments

The authors thank Dr. K.S. Nandakumar for critically reviewing and helping in writing the manuscript.

## References

1. S. F. Pedersen, Y.-C. Ho, et al., The Journal of clinical investigation 130, 2202 (2020).

2. E. Dong, H. Du, L. Gardner, The Lancet infectious diseases 20, 533 (2020).

3. Coronavirus cases: worldometers.info/coronavirus/.

4. Ü. Açikgöz, A. Günay, Turkish journal of medical sciences 50, 520 (2020).

5. A. Zaremba, D. Y. Aharon, E. Demir, R. Kizys, D. Zawadka, Research in International Business and Finance 56, 101359 (2021).

6. D. R. Q. Lemos, et al., Revista da Sociedade Brasileira de Medicina Tropical 53 (2020).

7. Z. Andreadakis, et al., Nature reviews. Drug discovery 19, 305 (2020).

8. M. Voysey, et al., The Lancet 397, 99 (2021).

9. A. van Dorn, The Lancet 396, 523 (2020).

10. D. Skegg, et al., The Lancet 397, 777 (2021).

11. S. A. Plotkin, Nature medicine 11, S5 (2005).

12. W. A. Orenstein, et al., Bulletin of the World Health Organization 63, 1055 (1985).

13. S. Piantadosi, Clinical trials: a methodologic perspective (John Wiley & Sons, 2017).

14. S. J. Pocock, Clinical trials: a practical approach (John Wiley & Sons, 2013).

15. R. DerSimonian, N. Laird, Controlled clinical trials 7, 177 (1986).

16. D. V. Mehrotra, et al., Annals of internal medicine 174, 221 (2021).

17. Y.-F. Tu, et al., International journal of molecular sciences 21, 2657 (2020).

18. S. J. Pocock, Blinding and Placebos (John Wiley and Sons, Ltd, 2013), chap. 6, pp. 90–99.

19. P. Olliaro, E. Torreele, M. Vaillant, The Lancet Microbe (2021).

20. S. J. Pocock, The Size of a Clinical Trial (John Wiley and Sons, Ltd, 2013), chap. 9, pp. 123–141.

21. E. Callaway, Nature.

22. L. J. Abu-Raddad, H. Chemaitelly, A. A. Butt, New England Journal of Medicine (2021).

23. A. Fontanet, et al., The Lancet 397, 952 (2021).

24. S. Elbe, G. Buckland-Merrett, Global challenges 1, 33 (2017).

25. M. Voysey, et al., The Lancet 397, 881 (2021).

26. D. Delen, E. Eryarsoy, B. Davazdahemami, JMIR public health and surveillance 6, e19862 (2020).

27. K. Kupferschmidt, M. Wadman, Delta variant triggers new phase in the pandemic (2021).

28. E. J. Haas, et al., The Lancet 397, 1819 (2021).

29. G. Strang, G. Strang, G. Strang, G. Strang, Introduction to linear algebra, vol. 3 (Wellesley-Cambridge Press Wellesley, MA, 1993).

30. D. Duong, Alpha, beta, delta, gamma: What’s important to know about sars-cov-2 variants of concern? (2021).

31. E. C. Wall, et al., Lancet (London, England) (2021).

32. E. C. Wall, et al., The Lancet 397, 2331 (2021).

